# The association between statin and COVID-19 adverse outcomes: National COVID-19 cohort in South Korea

**DOI:** 10.1101/2021.07.30.21261329

**Authors:** Ronald Chow, Jihui Lee, Hyerim Noh, Jongseong Lee, Hyun Joon Shin, Young-Geun Choi

## Abstract

**Background:** There currently exists limited and conflicting clinical data on the use of statins amongst COVID-19 patients. Given the both paucity and lack of consensus among data on statin’s efficacy and safety amongst COVID-19 patients, the current guideline is to continue statin in COVID-19 patients, who have previously been treated with statins. The aim of this paper was to compare hospitalized patients with COVID-19 who did and did not receive statins, in terms of COVID-19 outcomes.

**Methods:** We conducted population-based retrospective study using South Korea’s nationwide healthcare database as of May 15 2020. We identified 4,349 patients hospitalized with COVID-19 and aged 40 years or older. The cohort entry was defined as the date of hospitalization. Statin users were individuals with inpatient and outpatient prescription records of statins in the 240 days before cohort entry, and non-users were those without such records during this period. Our primary outcome was a composite endpoint of all-cause death, intensive care unit (ICU) admission, mechanical ventilation use and cardiovascular outcomes (myocardial infarction (MI), transient cerebral ischemic attacks (TIA) or stroke). We conducted inverse probability of treatment weighting (IPTW)-adjusted logistic regression analysis to estimate odds ratio (OR) and corresponding 95% confidence intervals (CI), to compare outcomes between statin users and non-users.

**Findings:** 1,115 patients were statin users (mean age = 65.9 years; 60% female), and 3,234 were non-users (mean age = 58.3 years; 64% female). Statin use was not associated with increased risk of the primary outcome (IPTW OR 0.82; 95% CI: 0.60-1.11). Subgroup analysis showed a protective role of statins, for individuals with hypertension (IPTW OR 0.40; 95% CI: 0.23-0.69, p for interaction: 0.0087).

**Interpretation:** Given that statins are not detrimental and that it may be beneficial amongst hypertensive patients and relatively cheap, we would encourage further investigation into statin for the prevention and treatment of COVID-19.

**Funding:** YGC’s work was partially supported by 2020R1G1A1A01006229 awarded by the National Research Foundation of Korea.

**Research in context:** *Evidence before this study:* There is limited and conflicting data reporting on statin use among COVID-19 patients, and its association with COVID-19 outcomes

*Added value of this study:* We report no difference in COVID-19 outcomes between patients who used and did not use statins prior to COVID-19 diagnosis, except in hypertensive patients in which statins was shown to have a protective effect.

*Implications of all the available evidence:* As statins are not detrimental and relatively cheap, we encourage further investigation into statin for the prevention and treatment of COVID-19.

## Introduction

On March 11, 2020, the World Health Organization (WHO) declared the COVID-19 outbreak a global pandemic^1,2^. Since then, the daily number of global COVID-19 cases has increased from a few thousand in the beginning of March, to 50,000 at the end of March, to 100,000 in May, and to 200,000 in July^3^.

The pathophysiology of the severe acute respiratory syndrome coronavirus 2 (SARS-CoV-2), which causes the COVID-19 disease, involves an overproduction of an early response proinflammatory cytokines, specifically tumour necrosis factor (TNF), IL-6 and IL-1β)^4^. If unabated, this cytokine storm subsequently places COVID-19 patients at increased risk of vascular hyperpermeability, multiorgan failure, and death^5^.

As a result, statins have been suggested for use as a therapy for COVID-19. It is reported that statin can stabilize MYD88 at normal levels and reduce an ensuing cytokine storm^6^. Statins may also up-regulate ACE2, which is typically downregulated by SARS-CoV-2 and facilitates the infiltration of SARS-CoV-2^7^. Statins can cause side effects of myalgia, increased creatine phosphokinase, and rhabdomyolysis and corresponding acute kidney injury, albeit quite rare; all these are reversible upon discontinuation of statins^8^.

There currently exists limited and conflicting clinical data on the use of statins amongst COVID-19 patients. While Zhang *et al*^9^ and Rodriguez-Nava *et al*^10^ report that statin reduced the risk of mortality and/or disease severity, and Wang *et al*^11^ reported an increased mortality among statin users, other studies^12-18^ report no significant difference between statin users and non-users. Meanwhile, a recent systematic review and meta-analysis of 110,078 patients reported a reduced risk of mortality among those administered statins after their COVID-19 hospitalization, but no difference in patients who were administered statins before hospitalization^19^. Although there is a lack of consensus among data on statin’s efficacy and safety amongst COVID-19 patients, the current recommendation is for COVID-19 patients to continue any antecedent statin use^20^.

Given the limited and conflicting data, there is a need for further investigation into the relationship between statin use and COVID-19 outcomes. The aim of this paper was to compare hospitalized patients with COVID-19 who did and did not receive statins, in terms of COVID-19 outcomes.

## Methods

### Data Source

We conducted a population-based cohort study using the Health Insurance Review and Assessment Service (HIRA) claims data released by the South Korean government. The HIRA is a sole nationwide governmental agency providing health insurance claims review, covering 98% of the Korean population (about 50 million), and employs a fee-for-service reimbursement system^21^. In March 2020, the HIRA released its claim database for all COVID-19 patients in South Korea. This population-level claims dataset includes information on sociodemographic characteristics, healthcare utilization history, diagnosis results (International Classification of Diseases, 10^th^ Revision^22^; ICD-10) and prescription from both inpatient and outpatient settings. COVID-19 diagnosis was determined through the South Korean Disease Control and Prevention Agency (KDCA) database. This database is the first nationwide dataset of COVID-19 patients. Details of the HIRA database are provided in Supplementary Material 1.

This study was approved by the Human Investigation Review Board of Public Institutional Bioethics Committee designated by the South Korean Ministry of Health and Welfare, which waived the requirement of informed consent due to retrospective study design and anonymity of the HIRA database (IRB # P01-2020-1262-001).

### Study Design and Participants

Between January 1, 2020 and May 15, 2020, 7,590 individuals tested positive for COVID-19. A positive COVID-19 result was defined as a positive result from South Korean Ministry of Food and Drug Safety–approved diagnostic tests that used a reverse-transcription polymerase chain reaction method^23^. Individuals were excluded if they were younger than 40 years old, since younger patients are less likely to experience COVID-19 outcomes regardless of statin use. As well, we excluded individuals who were not hospitalized (Figure 1), for precise outcome assessment. As a result, 4,349 individuals were included in the sample cohort. Cohort entry was defined as the date of admission for COVID-19 hospitalization. Most of these patients were hospitalized until they fully recovered. A full recovery was defined as cessation of fever without medication use, and 2 consecutive negative test results within a 24 hour period^24^.

**Figure 1.**
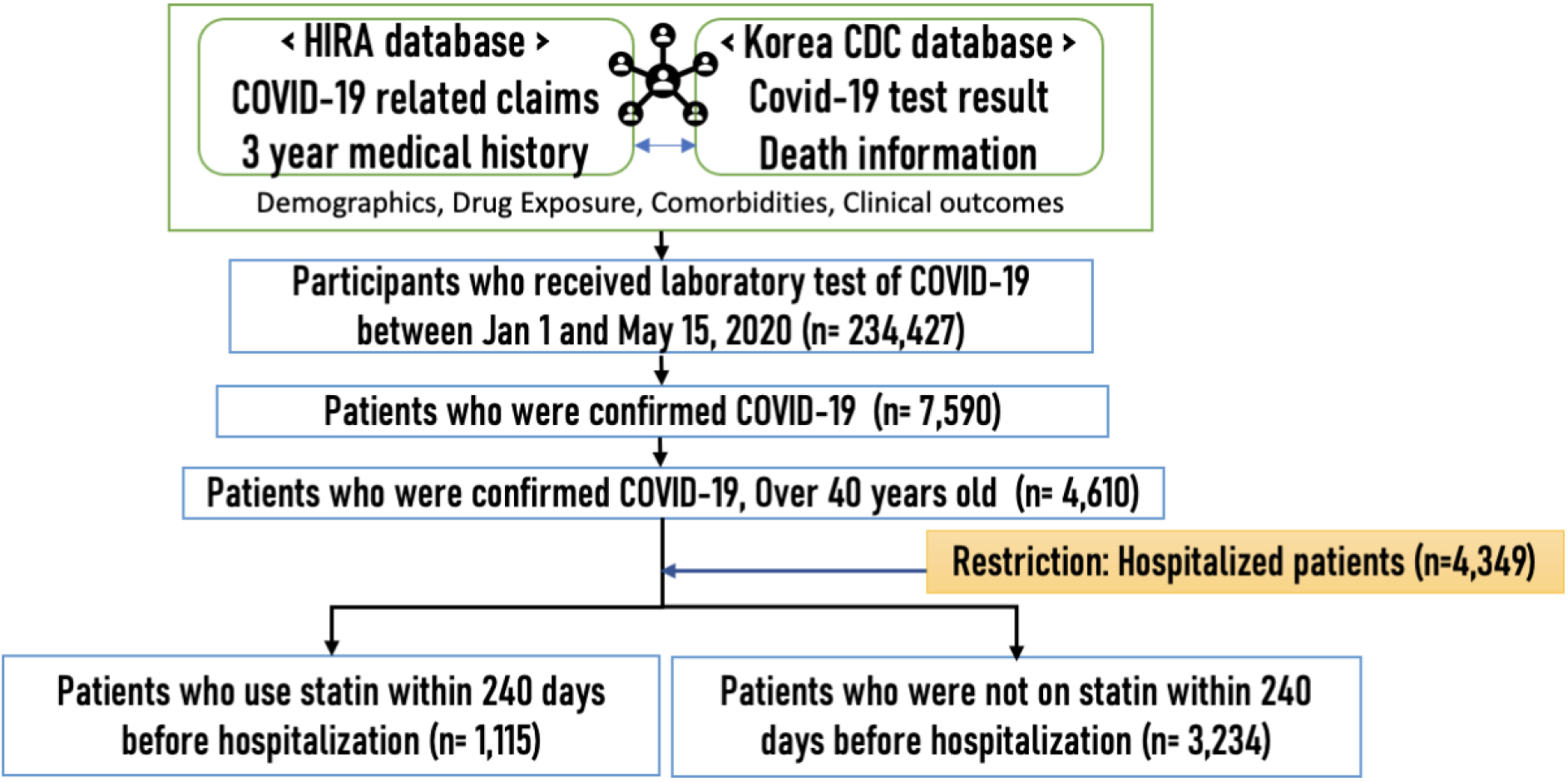
Population-based cohort study design using the HIRA and KDCA database of South Korea HIRA = Health Insurance Review and Assessment Service; KDCA = Korean Disease Control and Prevention Agency

### Statin Exposure

We defined exposure by using inpatient and outpatient prescription records of statin from the HIRA database (Supplementary Material 2). Our definition follows an intention-to-treat approach. Patients prescribed statin within 240 days prior to cohort entry were regarded as statin users, and otherwise defined as non-users.

### Outcomes

We set the primary outcome as a composite endpoint of all-cause death, intensive care unit (ICU) admission, mechanical ventilation use and cardiovascular outcomes (myocardial infarction (MI), transient cerebral ischemic attacks (TIA) or stroke). Secondary outcomes were the individual components of the composite endpoint. These outcomes were defined using in-hospital ICD-10 diagnostic codes and the national procedure coding system (Supplementary Material 2). We measured study outcomes between the date of cohort entry and the earliest of the date of hospital discharge or end of study period (May 15, 2020), whichever occurred earlier.

### Potential Confounders

We included sociodemographic and clinical factors that are considered to be associated with both statin use and risk of the outcomes. For sociodemographic factors, we included age in years, square of age, sex, and health insurance type at cohort entry. We included clinical variables of 20 comorbidities and use of 12 co-medications. The ascertainment period to define comorbidities was from 3 years prior (−1080d) through the start of the exposure ascertainment (−240d). The ascertainment period to define co-medications was from 2 years prior (−720d) through −240d. We defined comorbidity variables from in-hospital ICD-10 diagnostic codes and co-medications from in/out-hospital anatomical therapeutic chemical codes (Supplementary Material 2). When defining malignancy, we additionally used the expanded benefit codes in addition to diagnosis codes to reduce false-positives.

### Statistical Analyses

We summarized the baseline characteristics for statin users and non-users using mean and standard deviation for continuous variables, and frequency and percentage for categorical variables. We investigated imbalance of distribution of covariates between statin users and non-users using absolute standardized difference (aSD). An aSD ≤ 0.1 was preferred for balance.

We employed the inverse probability of treatment weighting (IPTW)-adjustment approach in order to reduce the confounding bias of statin users. Propensity score (PS) was estimated by a logistic regression with baseline sociodemographic and clinical characteristics as predictors (Table 1). We assigned the IPTW weight to each individual in the cohort, 1/PS to the statin users, and 1/(1-PS) to the non-users and constructed a balanced pseudo-population. Weighted univariable logistic regression models were fitted to estimate the effect of statin use on primary and secondary outcomes. For comparison, we also considered unweighted logistic regression: both univariable and multivariable, adjusted for age, sex, insurance type, history of diabetes and history of hypertension. For each logistic regression model, we reported the estimated odds ratio (OR) and its 95% confidence interval (CI).

**Table 1.** Baseline sociodemographic and clinical characteristics of COVID-19 patients with hospitalization >=40 years of age between January 1, 2020 and May 15, 2020 in South Korea.

|  | Before IPTW |  |  | After IPTW<sup>*</sup> |  |  |
| --- | --- | --- | --- | --- | --- | --- |
|  | Statin User*<br>1115 | Non-User<br>3234 | aSD | Statin User<br>4099 | Non-User<br>4505 | aSD |
| **Age (years; mean+sd)** | 65.9±11.2 | 58.3±12.3 | 0.64 | 61.1±13.2 | 60.9±12.9 | 0.01 |
| 40-49 | 79 (7.1) | 829 (25.6) |  | 892 (21.8) | 893 (19.8) |  |
| 50-59 | 252 (22.6) | 1155 (35.7) |  | 1080 (26.3) | 1432 (31.8) |  |
| 60-69 | 379 (34.0) | 684 (21.2) |  | 1097 (26.8) | 1066 (23.7) |  |
| 70-79 | 259 (23.2) | 312 (9.6) |  | 549 (13.4) | 644 (14.3) |  |
| 80-89 | 131 (11.7) | 202 (6.2) |  | 415 (10.1) | 380 (8.4) |  |
| 90+ | 15 (1.3) | 52 (1.6) |  | 67 (1.6) | 90 (2.0) |  |
| **Sex** |  |  |  |  |  |  |
| Male | 442 (39.6) | 1167 (36.1) | 0.07 | 1703 (41.5) | 1673 (37.1) | 0.09 |
| Female | 673 (60.4) | 2067 (63.9) | 0.07 | 2397 (58.5) | 2832 (62.9) | 0.09 |
| **Health insurance type** |  |  |  |  |  |  |
| Medical insurance | 960 (86.1) | 2871 (88.8) | 0.08 | 3490 (85.1) | 3952 (87.7) | 0.08 |
| Medical aid | 155 (13.9) | 363 (11.2) | 0.08 | 610 (14.9) | 553 (12.3) | 0.08 |
| **Comorbidities** |  |  |  |  |  |  |
| Arrhythmia | 46 (4.1) | 62 (1.9) | 0.13 | 112 (2.7) | 116 (2.6) | 0.01 |
| Asthma | 144 (12.9) | 279 (8.6) | 0.14 | 400 (9.8) | 458 (10.2) | 0.01 |
| Atrial fibrillation | 36 (3.2) | 35 (1.1) | 0.15 | 63 (1.5) | 72 (1.6) | 0.00 |
| Autoimmune disease | 117 (10.5) | 272 (8.4) | 0.07 | 303 (7.4) | 396 (8.8) | 0.05 |
| Chronic lung disease | 564 (50.6) | 794 (24.6) | 0.56 | 1425 (34.8) | 1487 (33.0) | 0.04 |
| Coronary artery disease | 187 (16.8) | 118 (3.6) | 0.44 | 320 (7.8) | 349 (7.7) | 0.00 |
| Dementia | 98 (8.8) | 189 (5.8) | 0.11 | 371 (9.1) | 330 (7.3) | 0.06 |
| Diabetes mellitus | 449 (40.3) | 345 (10.7) | 0.72 | 910 (22.2) | 933 (20.7) | 0.04 |
| Heart failure | 87 (7.8) | 90 (2.8) | 0.23 | 200 (4.9) | 213 (4.7) | 0.01 |
| Hyperlipidemia | 729 (65.4) | 585 (18.1) | 1.09 | 1335 (32.6) | 1480 (32.9) | 0.01 |
| Hypertension | 596 (53.5) | 571 (17.7) | 0.81 | 1197 (29.2) | 1332 (29.6) | 0.01 |
| Kidney disease | 28 (2.5) | 28 (0.9) | 0.13 | 65 (1.6) | 71 (1.6) | 0.00 |
| Liver disease | 44 (3.9) | 146 (4.5) | 0.03 | 295 (7.2) | 214 (4.8) | 0.10 |
| Malignancy | 61 (5.5) | 146 (4.5) | 0.04 | 223 (5.4) | 248 (5.5) | 0.00 |
| Other cerebrovascular diseases | 112 (10.0) | 125 (3.9) | 0.24 | 252 (6.1) | 279 (6.2) | 0.00 |
| Peripheral vascular disease | 144 (12.9) | 207 (6.4) | 0.22 | 398 (9.7) | 368 (8.2) | 0.05 |
| Pneumonia including tuberculosis | 95 (8.5) | 204 (6.3) | 0.08 | 411 (10.0) | 339 (7.5) | 0.09 |
| Psychiatric disorders | 357 (32.0) | 690 (21.3) | 0.24 | 1197 (29.2) | 1132 (25.1) | 0.09 |
| Stroke or TIA | 156 (14.0) | 129 (4.0) | 0.36 | 324 (7.9) | 350 (7.8) | 0.00 |
| Thromboembolism | 120 (10.8) | 93 (2.9) | 0.32 | 246 (6.0) | 248 (5.5) | 0.02 |
| **Co-medications** |  |  |  |  |  |  |
| Acetaminophen | 610 (54.7) | 1475 (45.6) | 0.18 | 1970 (48.1) | 2210 (49.1) | 0.02 |
| Antibiotics systemic | 840 (75.3) | 2167 (67.0) | 0.18 | 2778 (67.8) | 3114 (69.1) | 0.03 |
| Anticoagulants | 93 (8.3) | 76 (2.4) | 0.27 | 173 (4.2) | 207 (4.6) | 0.02 |
| Antidementia | 146 (13.1) | 263 (8.1) | 0.16 | 490 (12.0) | 457 (10.2) | 0.06 |
| Antidepressants | 199 (17.8) | 361 (11.2) | 0.19 | 667 (16.3) | 620 (13.8) | 0.07 |
| Antidiabetics | 397 (35.6) | 207 (6.4) | 0.77 | 647 (15.8) | 727 (16.1) | 0.01 |
| Antiplatelets | 420 (37.7) | 254 (7.9) | 0.76 | 722 (17.6) | 842 (18.7) | 0.03 |
| Antipsychotics | 269 (24.1) | 682 (21.1) | 0.07 | 1100 (26.8) | 1037 (23.0) | 0.09 |
| Antivirals | 79 (7.1) | 223 (6.9) | 0.01 | 284 (6.9) | 298 (6.6) | 0.01 |
| Anxiolytics | 390 (35.0) | 777 (24.0) | 0.24 | 1329 (32.4) | 1213 (26.9) | 0.12 |
| Immunosuppressant | 630 (56.5) | 1524 (47.1) | 0.19 | 2061 (50.3) | 2255 (50.1) | 0.00 |
| NSAIDs | 917 (82.2) | 2437 (75.4) | 0.17 | 3091 (75.4) | 3468 (77.0) | 0.04 |

aSD = absolute standardized difference; IPTW = inverse probability of treatment weighting; TIA = transient cerebral ischemic attack; NSAIDs = nonsteroidal anti-inflammatory drugs.
Mean and standard deviation were reported for continuous variables. Frequency and percentage were reported for categorical variables.
\*Patients prescribed within 240 days prior to cohort entry were regarded as statin users, and otherwise defined as non-users.
§Weighted cohort using the inverse probability of treatment weighting (IPTW).

### Subgroup Analyses

We stratified the cohort by (1) age (<65 and ≥ 65 years) and (2) sex for the risk of the primary outcome. We also considered stratification by the history of (3) hyperlipidemia, (4) hypertension, and (5) diabetes mellitus. For each subgroup analysis, we recalculated PS by multivariable logistic regression. We obtained p for interaction by each subgroup.

### Sensitivity Analyses

We conducted sensitivity analyses by extending the scope of the study population in two settings. The first setting was to redefine the study population by relaxing the hospitalization condition. That is, we included all confirmed positive individuals of at least 40 years of age regardless of their hospitalization record. The second setting was to extend the time window for exposure. To capture the patients who infrequently received statin prescription, we extended the time window for ascertaining exposure from 240 to 360 days, prior to cohort entry.

To investigate the sensitivity of results against the choices of statistical methods, we considered the following alternative approaches. First, we repeated the main IPTW analysis with discarding individuals with extreme PS values, PS < 0.025 or PS > 0.975 (IPTW with trimming). Second, in our unweighted multivariable logistic regression models, we included the estimated PSs as an additional predictor to other covariates (outcome adjustment model). Third, we applied the stabilized mortality ratio weighting, 1 for the user group and PS/(1-PS) for the non-user group, instead of IPTW (SMR weighting). Lastly, we considered propensity score matching (PS matching).

A two-sided p-value less than 0.05 was considered statistically significant. All analyses were conducted using R 3.5.2 (R Core Team, Vienna, Austria).

## Results

A total of 4,349 adults of at least 40 years of age were hospitalized with COVID-19 in South Korea between January 1, 2020 and May 15, 2020 and included this analysis. Of these, 1,115 (27%) were statin users. Statin users were older than non-users on average (65.9 ± 11.2 vs 58.3 ± 12.3). Among statin users, 39.6% of statin users were males, whereas 36.1% of non-users were male. A larger proportion of statin users had comorbidities of chronic lung disease (50.6% vs 24.6%), diabetes mellitus (40.3% vs 10.7%), hyperlipidemia (65.4% vs 18.1%), and hypertension (53.5% vs 17.7%), relative to non-users. Statin users also used more antidiabetic (35.6% vs 6.4%) and antiplatelet (37.7% vs 7.9%) medications (Table 1).

Among 530 primary composite events of all-cause death, ICU admission, mechanical ventilation use and cardiovascular adverse outcomes, 362 occurred in non-users (11.2%; 362/3234) and 168 (15.1%; 168/1115) in statin users. No difference was noted between statin users and non-users in the IPTW weighted analysis (IPTW adjusted OR 0.82; 95% CI: 0.60-1.11). Similarly, there was no difference between statin users and non-users for the individual components of all-cause death, mechanical ventilation, ICU admission, and cardiovascular adverse outcome (Table 2).

**Table 2.** Risk of adverse clinical outcomes associated with statin use among COVID-19 >= 40 years of age with hospitalization

|  | Number<br>of<br>Patients | Number<br>of<br>Events | Cumulative<br>Incidence (%) | Odds Ratio (95% Confidence Interval) |  |  |
| --- | --- | --- | --- | --- | --- | --- |
|  |  |  |  | Unadjusted* | Adjusted<sup>§</sup> | IPTW Weighted<sup>#</sup> |
| **All-cause death, ICU admission, mechanical ventilation, MI, stroke, TIA** |  |  |  |  |  |  |
| Non-user | 3234 | 362 | 11.2 | 1.00 (reference) | 1.00 (reference) | 1.00 (reference) |
| User | 1115 | 168 | 15.1 | 1.41 (1.16-1.71) | 0.87 (0.70-1.09) | 0.82 (0.60-1.11) |
| **All-cause death** |  |  |  |  |  |  |
| Non-user | 3234 | 131 | 4.1 | 1.00 (reference) | 1.00 (reference) | 1.00 (reference) |
| User | 1115 | 86 | 7.7 | 1.98 (1.50-2.62) | 0.97 (0.70-1.34) | 0.90 (0.58-1.39) |
| **Mechanical ventilation** |  |  |  |  |  |  |
| Non-user | 3234 | 66 | 2.0 | 1.00 (reference) | 1.00 (reference) | 1.00 (reference) |
| User | 1115 | 57 | 5.1 | 2.59 (1.80-3.71) | 1.41 (0.94-2.09) | 1.24 (0.74-2.09) |
| **ICU admission** |  |  |  |  |  |  |
| Non-user | 3234 | 233 | 7.2 | 1.00 (reference) | 1.00 (reference) | 1.00 (reference) |
| User | 1115 | 97 | 8.7 | 1.23 (0.96-1.57) | 0.97 (0.73-1.28) | 0.72 (0.49-1.05) |
| **Cardiovascular adverse outcome (MI, stroke and TIA)** |  |  |  |  |  |  |
| Non-user | 3234 | 31 | 1.0 | 1.00 (reference) | 1.00 (reference) | 1.00 (reference) |
| User | 1115 | 15 | 1.3 | 1.41 (0.76-2.62) | 1.17 (0.60-2.29) | 0.79 (0.32-1.97) |

MI = myocardial infarction; TIA = transient cerebral ischemic attack.
\* Unweighted univariable logistic regression model
§ Unweighted multivariable logistic regression model adjusted for age, sex, insurance type, history of diabetes and history of hypertension

In the subgroup analyses, there existed a significant effect modification by hypertension status (*p* = 0.0087). Statin users who had hypertension experienced lower odds for the composite (IPTW OR 0.40, 95% CI: 0.23 – 0.69). There was no effect modification by age, sex, hyperlipidemia and diabetes mellitus (Figure 2).

**Figure 2.**
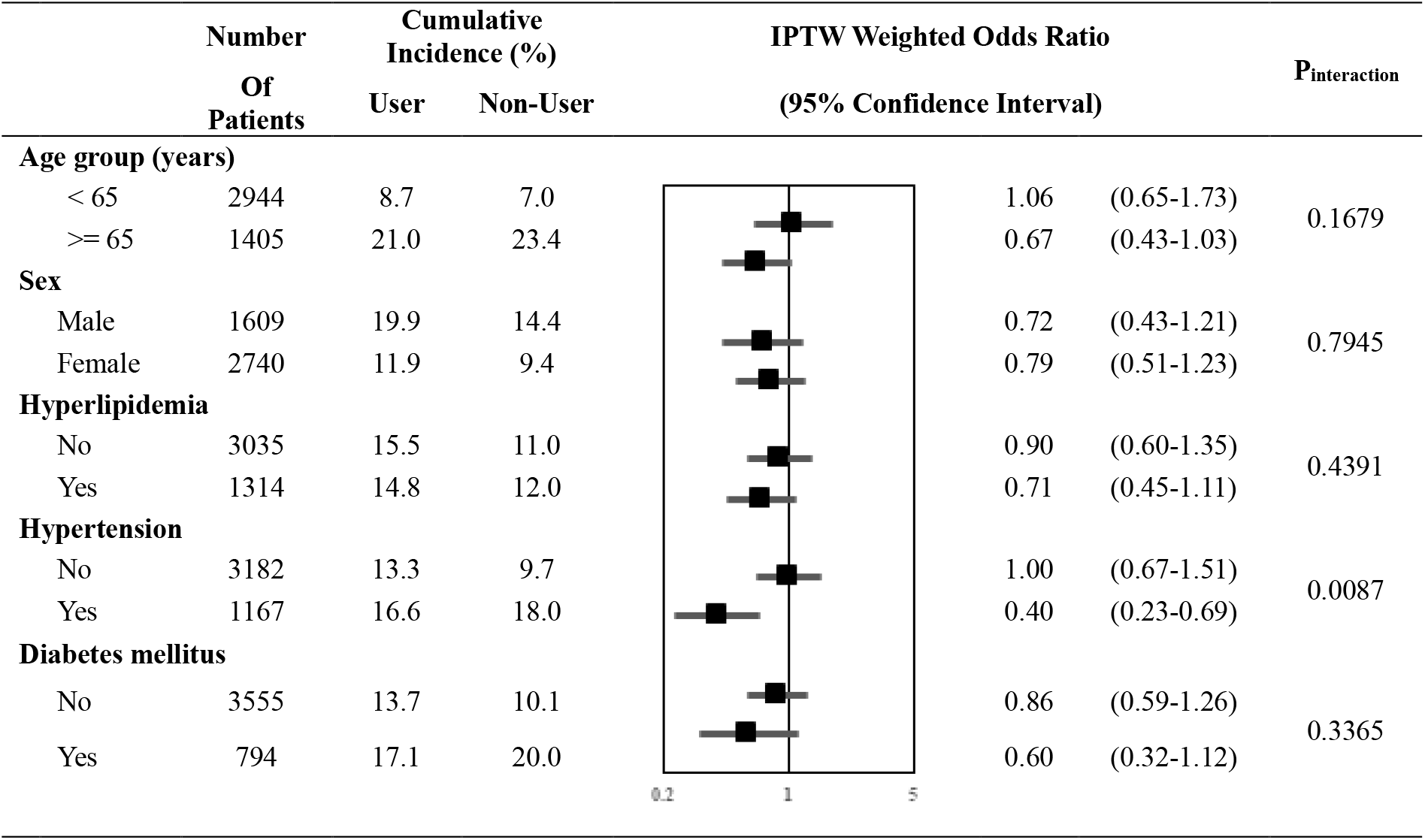
Forest plot summarizing the risk of primary outcome associated with statin use when stratified for age, sex, history of hyperlipidaemia, hypertension, and diabetes mellitus.

The results from the sensitivity analyses remained generally consistent with the main analysis with no harmful association between statin use and the outcomes. When redefining the study population, changing the exposure ascertainment window and applying other statistical methods, the primary outcome, all-cause death, ICU admission and cardiovascular outcomes showed generally similar results. Statins showed a protective association with the primary outcome, when using SMR weighting (OR 0.64, 95% CI: 0.45 – 0.93) and PS matching (OR 0.74, 95% CI: 0.56 – 0.98). For all-cause deaths, statins showed protective association when using SMR weighting (OR 0.56, 95% CI: 0.35 – 0.89) and PS matching (OR 0.67, 95% CI: 0.46 – 0.98). As well, statins showed a protective association for all-cause death, when including all confirmed patients with COVID-19 (OR 0.81, 95% CI: 0.68 – 0.96). For ICU admission, statin showed protective association when using IPTW weighting with trimming (IPTW OR 0.67, 95% CI: 0.47 – 0.97), or redefining exposure window to 360 days (IPTW OR 0.67, 95% CI: 0.47 – 0.97) (Supplementary Material 3-7).

## Discussion

This study reported on one of the largest investigations of COVID-19 patients with hospitalization in South Korea. It utilized a nationwide and completely enumerated dataset, and applied a propensity score-based weighting to control for potential confounding variables. In the main analysis, there was no significant difference between baseline (pre-hospitalization) statin use vs non-use, with respect to the outcomes of mortality, ICU admission, mechanical ventilation use and cardiovascular outcomes.

Even though our main analysis results showed null association between statin and all cause death, our sensitivity analysis of all cause death showed protective association with statin when using SMR weighting, PS matching or when including all confirmed patients with COVID-19. This finding is similar to report by Lee *et al*^25^, which showed a protective association of statins for all cause death in the similar cohort while using PS matching with a several different methodological choices from our study (first, they included patients of all age, rather than restricting their analyses to patients who were 40 years old and older; second, statin exposure was defined by Lee *et al* as a prescription up to 90 days prior to COVID-19 diagnosis rather than 240 days prior to COVID-19 diagnosis; third, they used Cox survival analysis while we used logistic regression). It therefore is possible that we missed detecting real protective association of statin in the main analysis, especially since some of sensitivity analysis showed protective association of statin for primary outcome, all cause death and ICU admission based on varying statistical methods, changing the exposure window change to 360 days and inclusion of non-hospitalized COVID-19 patients.

Also, in the prespecified subgroup analysis, statin was found to be associated with lower odds of the primary endpoint in hypertensive patients. As hypertensive COVID-19 patients are known to have poorer prognosis than normotensive patients^26^, this result is encouraging; statins may have a greater protective effect, for this sicker cohort of patients with hypertension. Further investigation is needed since statins may be cost-effective, with the potential benefit outweighing any potential toxicities^27^. Mechanistically, statins is postulated to upregulate ACE2, countering the effect of SARS-CoV-2 where ACE2 is downregulated and thereby facilitates the infiltration of SARS-CoV-2^7^. Although statin can cause muscle and liver side effects, discontinuation of statins treatment is known reverse these side effects, alleviating concerns of significant and irreversible adverse effects.

This study has several strengths. First, we used a nationwide cohort of all hospitalized patients with COVID-19 in South Korea between January 1, 2020 and May 15, 2020. Since South Korea maintained a strict patient management system during this period, the use of this population-based cohort may mitigate any potential sampling bias issue. In addition, relative to the study by Lee *et al*^25^, our study design aimed to further mitigate healthy user bias, by including chronic co-medications as confounders, which may have adjusted for past adherence^28^. Furthermore, our main findings of no harmful association between statin and COVID-19 outcomes were supported by our sensitivity analyses which showed even protective association when we redefined study population, extended the ascertainment period of the statin use, and considered various statistical approaches. Given these neutral to protective association between statin and COVID-19 outcomes, our findings may support the current guideline which is to continue statin in COVID-19 patients, who have previously been treated with statins^20^.

There are also several limitations of this study. First, there may have been outcome misclassification in cardiovascular outcome (MI, stroke, and TIA), because we defined our cardiovascular outcome variables by diagnostic codes in admirative claim data. HIRA reported that 82% of primary diagnosis codes in claims coincide with electronic medical records,^29^ so more than 18% of outcome misclassification can occur while trying to capture all the diagnostic codes other than primary diagnosis codes. However, we expect a greater validity for MI, stroke, and TIA since our study population focused on hospitalized patients under careful management. Other outcomes such as all-cause death seem to be well classified because the HIRA COVID-19 database was linked to the national death records and outcomes defined from procedure codes (ICU admission and mechanical ventilation use) are also expected to be valid since the codes are mandatory in the reimbursement review process. Second, statin exposure was defined based on inpatient and outpatient prescription. We are not aware of detailed information on the exposure, for example, adherence to the prescription. Last, although we performed IPTW and thorough sensitivity analyses, due to the inherent limitation of claims data, there might be still a residual confounding by potential unmeasured confounders typically used in clinical studies (e.g., body mass index, baseline blood pressures, laboratory test values).

In conclusion, our analysis of a nationwide sample of South Korean, hospitalized, COVID-19 patients found that statin users overall did not experience poorer COVID-19 outcomes, relative to non-users. However, in hypertensive patients, a protective association between statin use and COVID-19 outcomes were shown, which calls for further investigation. Given that statins are relatively cheap and not detrimental in this setting, we would encourage further investigation into statin for the prevention and treatment of COVID-19.

## Data Availability

YGC and HJS conceived this project. YGC and HN had full access to data for analysis. RC, Jihui L and Jongseong L drafted the manuscript. All authors contributed significantly, edited the manuscript and are accountable for its reporting.

## Acknowledgements

The authors thank healthcare professionals dedicated to treating COVID-19 patients in South Korea, the Ministry of Health and Welfare, the Health Insurance Review and Assessment Service (HIRA), and Ye-Jin Sohn (HIRA) for sharing invaluable national health insurance claims data.

## Declaration of interest

All authors completed and submitted the ICMJE Form for Disclosure of Potential Conflicts of Interest. The authors declare no competing interests.

## Data sharing

No additional data available.

## Supplementary Material Legends

Supplementary Material 1. Data schema of the Health Insurance Review and Assessment Service database.

Supplementary Material 2. Diagnosis codes based on the Korean Standard Classification of Diseases, 7^th^ Revision or International Classification of Disease, 10^th^ Revision codes, National Procedure codes, and drug codes based on World Health Organization-Anatomical Therapeutic Chemical classification codes.

Supplementary Materials 3-7. The results of sensitivity analysis, by outcomes.

### Supplementary Material 1

Data schema of the Health Insurance Review and Assessment Service (HIRA) database

#### 1. Data Description

The database used in this study is the Health Insurance Review and Assessment Service (HIRA) claim data released by the South Korean government as the world’s first de-identified COVID-19 nationwide data. Since South Korea have implemented a single National Health Insurance system, this administrative data includes the entire health insurance claims and clinical information of South Korean population. Information on non-essential medical treatment that are not covered by NHI was not collected based on fee-for-service payment system.

#### 2. Data Scope

The list of patients of COVID-19 (using submitted claims data) is connected to their history of medical service use for the past 5 years (using finalized claims data, from January 2015 to February 2020), and the entire data set is de-identified. All benefit claims relevant to COVID-19 as of March 15, 2020.

#### 3. Data Extraction Criteria

The databased include all health insurance types such as National Health Insurance, Medical Aid, Korea veterans service. COVID19 related claims consists of i) claims with “3/02” in classification code of MT043 (Target of medical cost support due to national disaster) based on the serial number of claim statement, ii) claims that contained “D6584” COVID-19 (Real-time polymerase chain reaction), iii) claims that contained COVID-19 related disease code (B342, B972, Z208, Z290, U18, U181, Z038, Z115, U071, U072), and iv)·claims that contained other COVID-19 related fee code (COVID-19 related admission fee, management fee, IUD, isolation, public relief hospital, residential treatment center, screening center, negative pressure room, etc.).

#### 4. Data Schema

The data based consists of COVID-19 related claims data, Healthcare service use history of the individuals who submitted COVID-19 related claims for the past 3 years. COVID-19 confirmation information and some related results such as was extracted from KCDA database. The included information are;

- Sociodemographic characteristics (i.e., sex, age, medical institution type)
- Diagnosis information (i.e., ICD-10 diagnosis codes, main diagnosis, sub diagnosis)
- Healthcare utilization information (i.e., number of visit, date of visit, duration of hospital stay)
- Medical expenditure information (i.e., out of pocket medical expenditure, reimbursed expenditure from NHI)
- Prescription information (i.e., national drug chemical code, number of supply, date of supply, dosage)

**Note**: COVID-19, coronavirus disease 2019; ICD-10, International Classification of Disease 10^th^ Revision. Detailed information on data schema is available upon request.

### Supplementary Material 2

Diagnosis codes based on the Korean Standard Classification of Diseases, 7^th^ Revision or International Classification of Disease, 10^th^ Revision codes, National Procedure codes, and drug codes based on World Health Organization-Anatomical Therapeutic Chemical classification codes.

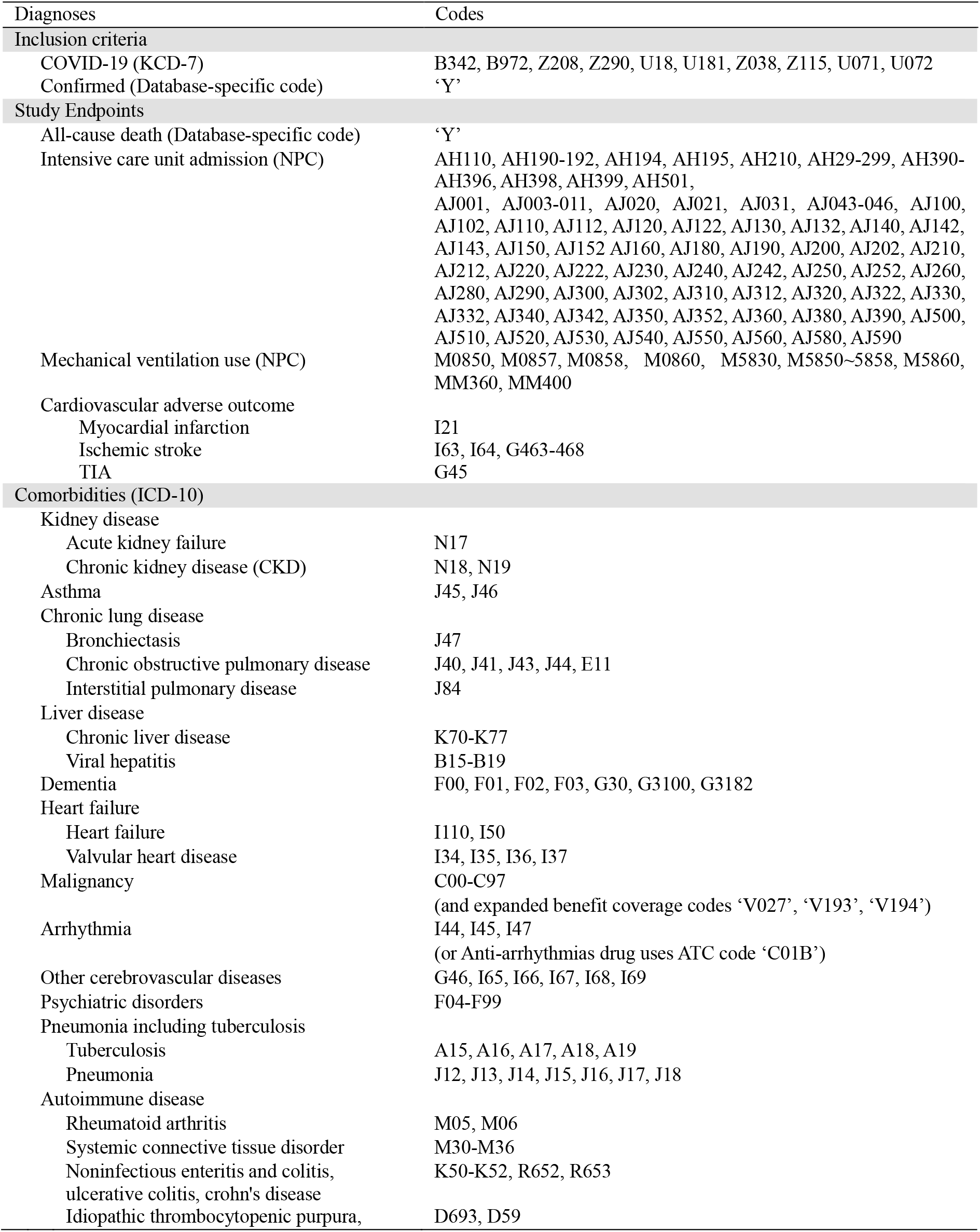
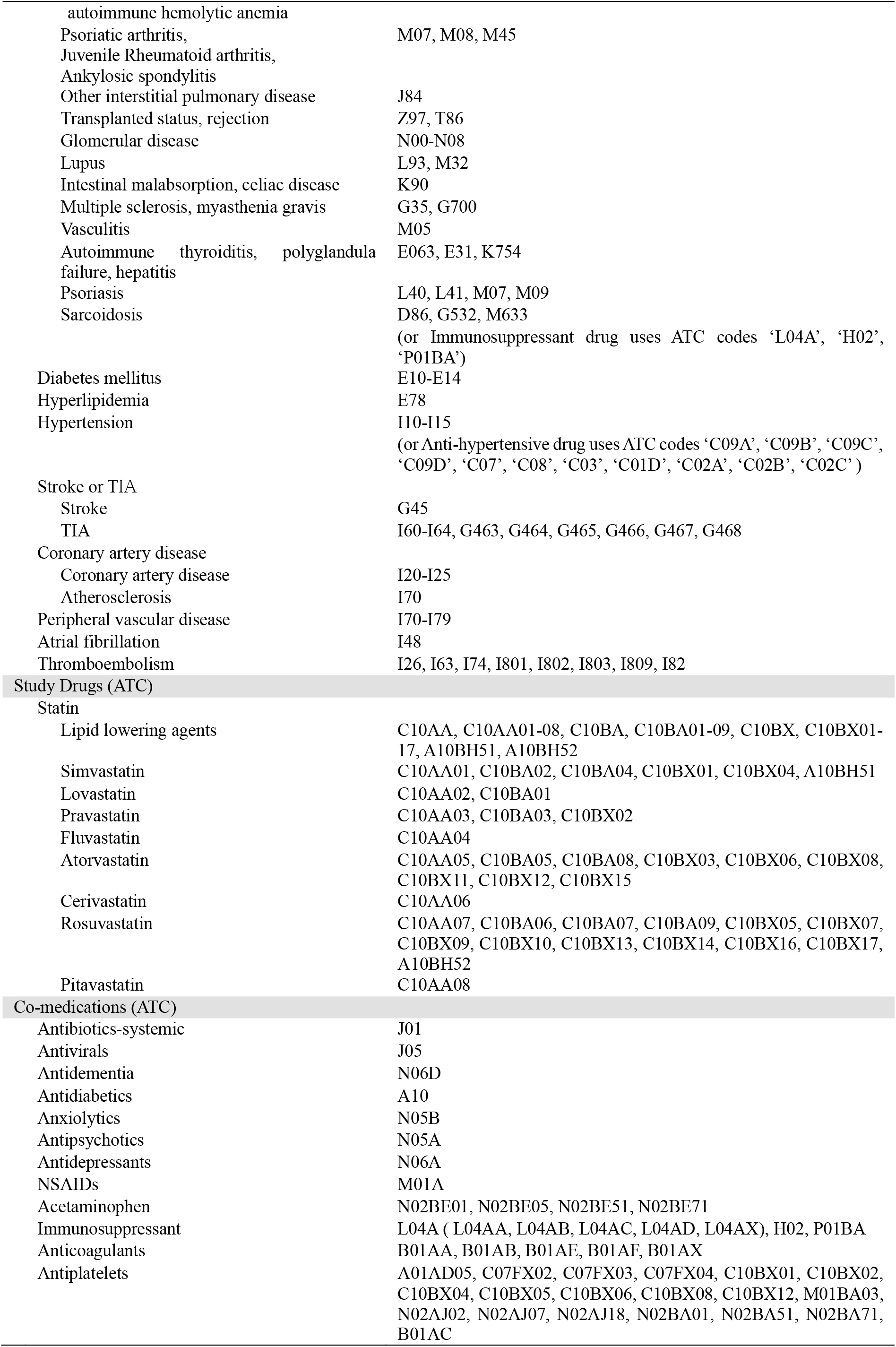
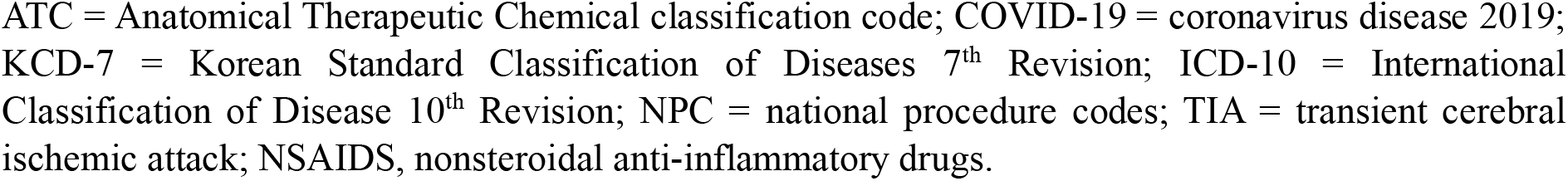

### Supplementary Material 3

Risk of primary outcome associated with statin use among COVID-19 patients with >= 40 years of age.

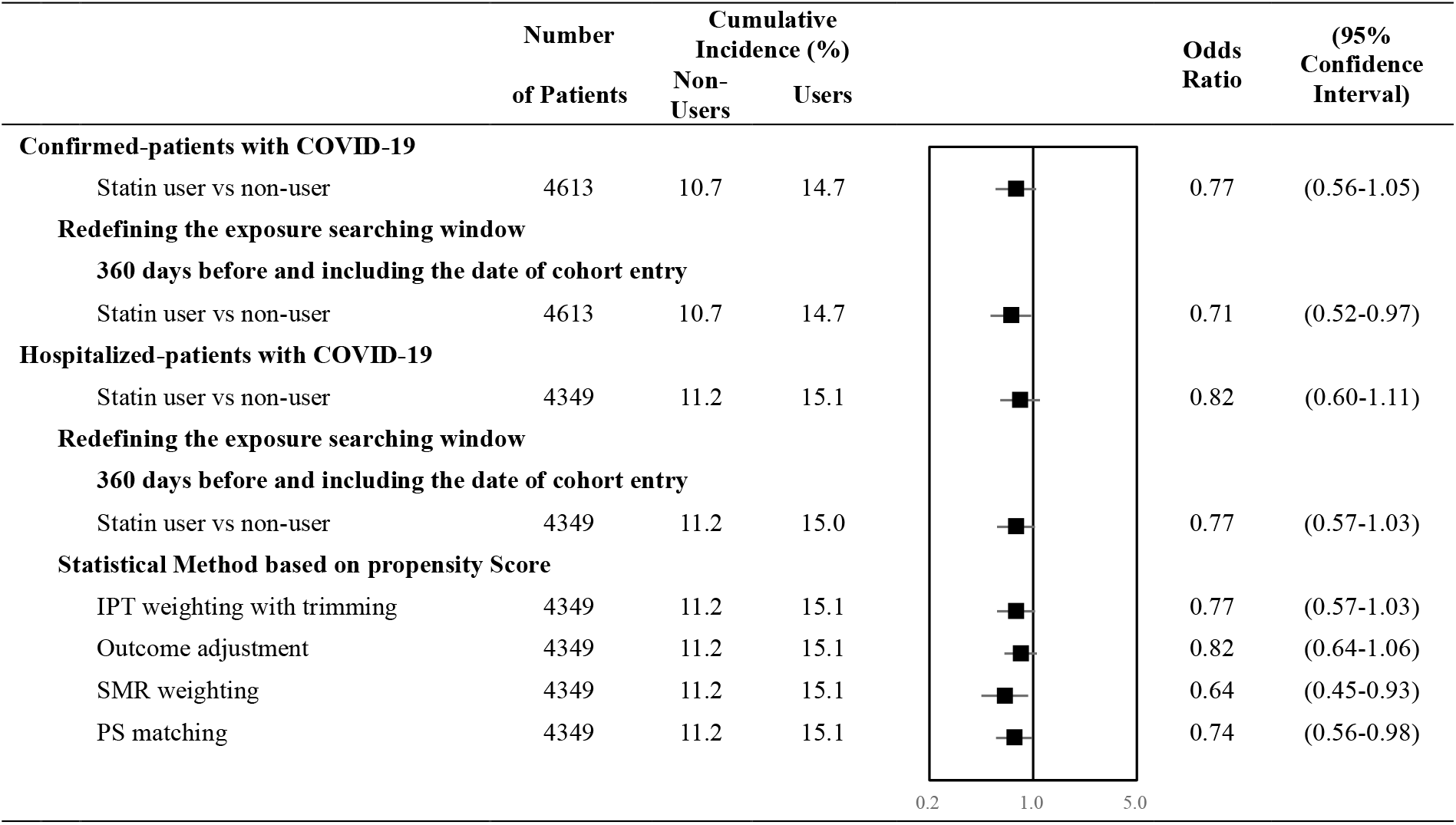

|  | Number<br>of Patients | Cumulative<br>Incidence (%) |  |  | Odds<br>Ratio | (95%<br>Confidence<br>Interval) |
| --- | --- | --- | --- | --- | --- | --- |
| **Confirmed-patients with COVID-19** |  |  |  |  |  |  |
| Statin user vs non-user | 4613 | 10.7 | 14.7 |  | 0.77 | (0.56-1.05) |
| **Redefining the exposure searching window** |  |  |  |  |  |  |
| **360 days before and including the date of cohort entry** |  |  |  |  |  |  |
| Statin user vs non-user | 4613 | 10.7 | 14.7 |  | 0.71 | (0.52-0.97) |
| **Hospitalized-patients with COVID-19** |  |  |  |  |  |  |
| Statin user vs non-user | 4349 | 11.2 | 15.1 |  | 0.82 | (0.60-1.11) |
| **Redefining the exposure searching window** |  |  |  |  |  |  |
| **360 days before and including the date of cohort entry** |  |  |  |  |  |  |
| Statin user vs non-user | 4349 | 11.2 | 15.0 |  | 0.77 | (0.57-1.03) |
| **Statistical Method based on propensity Score** |  |  |  |  |  |  |
| IPT weighting with trimming | 4349 | 11.2 | 15.1 |  | 0.77 | (0.57-1.03) |
| Outcome adjustment | 4349 | 11.2 | 15.1 |  | 0.82 | (0.64-1.06) |
| SMR weighting | 4349 | 11.2 | 15.1 |  | 0.64 | (0.45-0.93) |
| PS matching | 4349 | 11.2 | 15.1 |  | 0.74 | (0.56-0.98) |

### Supplementary Material 4

Risk of all-cause death associated with statin use among COVID-19 patients with >= 40 years of age.

|  | Number<br>of Patients | Cumulative<br>Incidence (%) |  |  | Odds<br>Ratio | (95%<br>Confidence<br>Interval) |
| --- | --- | --- | --- | --- | --- | --- |
|  |  | Non-Users | Users |  |  |  |
| **Confirmed-patients with COVID-19** |  |  |  |  |  |  |
| Statin user vs non-user | 4613 | 3.9 | 7.7 |  | 0.81 | (0.64-0.96) |
| **Redefining the exposure searching window** |  |  |  |  |  |  |
| **360 days before and including the date of cohort entry** |  |  |  |  |  |  |
| Statin user vs non-user | 4613 | 3.8 | 7.8 |  | 0.75 | (0.46-1.21) |
| **Hospitalized-patients with COVID-19** |  |  |  |  |  |  |
| Statin user vs non-user | 4349 | 4.1 | 7.7 |  | 0.90 | (0.58-1.39) |
| **Redefining the exposure searching window** |  |  |  |  |  |  |
| **360 days before and including the date of cohort entry** |  |  |  |  |  |  |
| Statin user vs non-user | 4349 | 4.0 | 7.8 |  | 0.87 | (0.57-1.33) |
| **Statistical Method based on Propensity Score** |  |  |  |  |  |  |
| IPT weighting with trimming | 4349 | 4.1 | 7.7 |  | 0.87 | (0.57-1.33) |
| Outcome adjustment | 4349 | 4.1 | 7.7 |  | 0.84 | (0.59-1.21) |
| SMR weighting | 4349 | 4.1 | 7.7 |  | 0.56 | (0.35-0.89) |
| PS matching | 4349 | 4.1 | 7.7 |  | 0.67 | (0.46-0.98) |

### Supplementary Material 5

Risk of adverse mechanical ventilation associated with statin use among COVID-19 patients with >= 40 years of age.

|  | Number<br>of Patients | Cumulative<br>Incidence (%) |  |  | Odds<br>Ratio | (95%<br>Confidence<br>Interval) |
| --- | --- | --- | --- | --- | --- | --- |
|  |  | Non-Users | Users |  |  |  |
| **Confirmed-patients with COVID-19** |  |  |  |  |  |  |
| Statin user vs non-user | 4613 | 1.9 | 4.8 |  | 1.23 | (0.73-2.06) |
| **Redefining the exposure searching window** |  |  |  |  |  |  |
| **360 days before and including the date of cohort entry** |  |  |  |  |  |  |
| Statin user vs non-user | 4613 | 1.9 | 4.7 |  | 1.18 | (0.72-1.95) |
| **Hospitalized-patients with COVID-19** |  |  |  |  |  |  |
| Statin user vs non-user | 4349 | 2.0 | 5.1 |  | 1.24 | (0.74-2.09) |
| **Redefining the exposure searching window** |  |  |  |  |  |  |
| **360 days before and including the date of cohort entry** |  |  |  |  |  |  |
| Statin user vs non-user | 4349 | 2.0 | 5.0 |  | 1.17 | (0.71-1.94) |
| **Statistical Method based on Propensity Score** |  |  |  |  |  |  |
| IPT weighting with trimming | 4349 | 2.0 | 5.1 |  | 1.18 | (0.71-1.95) |
| Outcome adjustment | 4349 | 2.0 | 5.1 |  | 1.22 | (0.77-1.95) |
| SMR weighting | 4349 | 2.0 | 5.1 |  | 1.09 | (0.63-1.90) |
| PS Matching | 4349 | 2.0 | 5.1 |  | 1.03 | (0.63-1.70) |

### Supplementary Material 6

Risk of ICU admission associated with statin use among COVID-19 patients with >=40 years of age

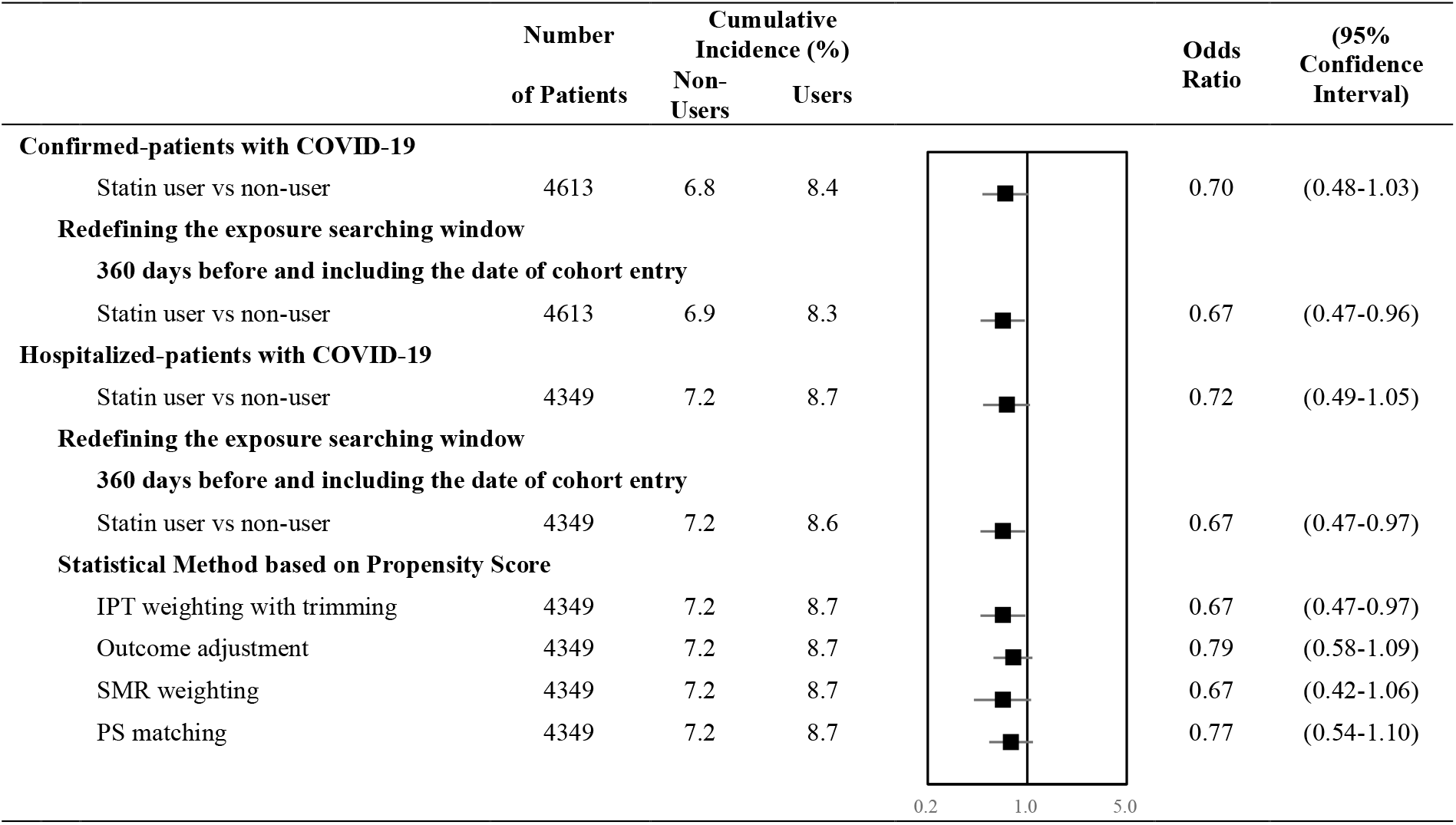

|  | Number<br>of Patients | Cumulative<br>Incidence (%) |  |  | Odds<br>Ratio | (95%<br>Confidence<br>Interval) |
| --- | --- | --- | --- | --- | --- | --- |
| **Confirmed-patients with COVID-19** |  |  |  |  |  |  |
| Statin user vs non-user | 4613 | 6.8 | 8.4 |  | 0.70 | (0.48-1.03) |
| **Redefining the exposure searching window** |  |  |  |  |  |  |
| **360 days before and including the date of cohort entry** |  |  |  |  |  |  |
| Statin user vs non-user | 4613 | 6.9 | 8.3 |  | 0.67 | (0.47-0.96) |
| **Hospitalized-patients with COVID-19** |  |  |  |  |  |  |
| Statin user vs non-user | 4349 | 7.2 | 8.7 |  | 0.72 | (0.49-1.05) |
| **Redefining the exposure searching window** |  |  |  |  |  |  |
| **360 days before and including the date of cohort entry** |  |  |  |  |  |  |
| Statin user vs non-user | 4349 | 7.2 | 8.6 |  | 0.67 | (0.47-0.97) |
| **Statistical Method based on Propensity Score** |  |  |  |  |  |  |
| IPT weighting with trimming | 4349 | 7.2 | 8.7 |  | 0.67 | (0.47-0.97) |
| Outcome adjustment | 4349 | 7.2 | 8.7 |  | 0.79 | (0.58-1.09) |
| SMR weighting | 4349 | 7.2 | 8.7 |  | 0.67 | (0.42-1.06) |
| PS matching | 4349 | 7.2 | 8.7 |  | 0.77 | (0.54-1.10) |

### Supplementary Material 7

Risk of cardiovascular outcomes (myocardial infarction, ischemic stroke and transient ischemic attack,) associated with statin use among COVID-19 patients with >= 40 years of age

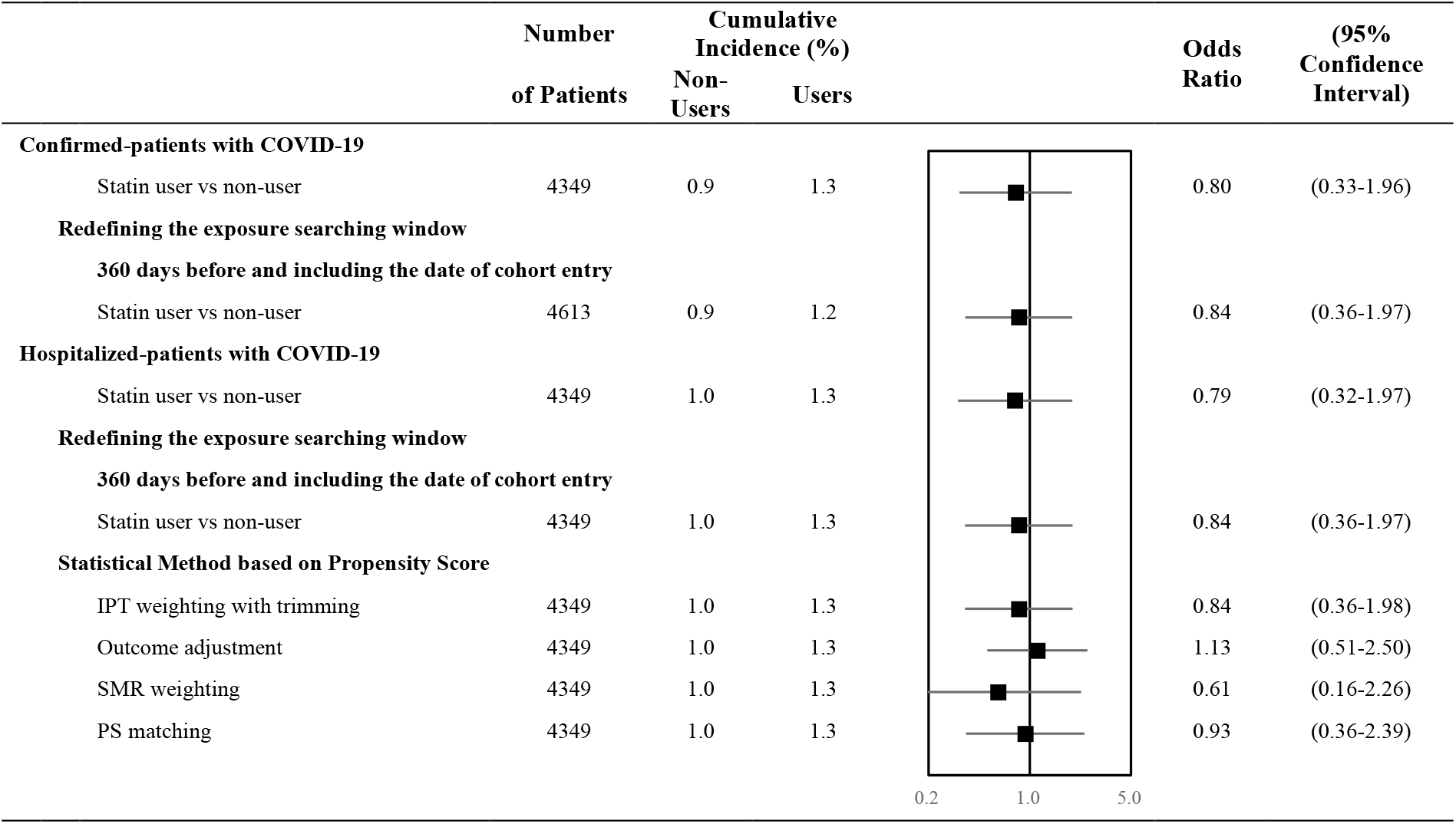

## Notes

### Competing Interest Statement

The authors have declared no competing interest.

### Funding Statement

YGC was partially supported by 2020R1G1A1A01006229 awarded by the National Research Foundation of Korea.

## References

1. Chow R, Elsayeed S, Lock M. How robust are the results of one of the first positive trials exploring hydroxycloroquine for treatment of COVID-19? medRxiv 2020. DOI: 10.1101/2020.05.06.20093237.

2. Chow R, Simone CB, 2nd, Lock M. Hydroxychloroquine for the treatment of COVID-19: the importance of scrutiny of positive trials. Ann Palliat Med 2020. DOI: 10.21037/apm-20-1538.

3. World Health Organizaion. Timeline: WHO’s COVID-19 response. World Health Organization. (https://www.who.int/emergencies/diseases/novel-coronavirus-2019/interactive-timeline#!).

4. Jose RJ, Manuel A. COVID-19 cytokine storm: the interplay between inflammation and coagulation. Lancet Respir Med 2020;8(6):e46–e47. DOI: 10.1016/s2213-2600(20)30216-2.

5. Meduri GU, Kohler G, Headley S, Tolley E, Stentz F, Postlethwaite A. Inflammatory cytokines in the BAL of patients with ARDS. Persistent elevation over time predicts poor outcome. Chest 1995;108(5):1303–14. DOI: 10.1378/chest.108.5.1303.

6. Dashti-Khavidaki S, Khalili H. Considerations for Statin Therapy in Patients with COVID-19. Pharmacotherapy 2020;40(5):484–486. DOI: 10.1002/phar.2397.

7. Fedson DS, Opal SM, Rordam OM. Hiding in Plain Sight: an Approach to Treating Patients with Severe COVID-19 Infection. mBio 2020;11(2). DOI: 10.1128/mBio.00398-20.

8. Guan WJ, Ni ZY, Hu Y, et al. Clinical Characteristics of Coronavirus Disease 2019 in China. N Engl J Med 2020;382(18):1708–1720. DOI: 10.1056/NEJMoa2002032.

9. Zhang XJ, Qin JJ, Cheng X, et al. In-Hospital Use of Statins Is Associated with a Reduced Risk of Mortality among Individuals with COVID-19. Cell Metab 2020;32(2):176-187.e4. DOI: 10.1016/j.cmet.2020.06.015.

10. Rodriguez-Nava G, Trelles-Garcia DP, Yanez-Bello MA, Chung CW, Trelles-Garcia VP, Friedman HJ. Atorvastatin associated with decreased hazard for death in COVID-19 patients admitted to an ICU: a retrospective cohort study. Crit Care 2020;24(1):429. DOI: 10.1186/s13054-020-03154-4.

11. Wang B, Van Oekelen O, Mouhieddine TH, et al. A tertiary center experience of multiple myeloma patients with COVID-19: lessons learned and the path forward. Journal of Hematology & Oncology 2020;94:13.

12. Argenziano MG, Bruce SL, Slater CL, et al. Characterization and clinical course of 1000 patients with coronavirus disease 2019 in New York: retrospective case series. BMJ 2020;369:m1996. DOI: 10.1136/bmj.m1996.

13. Ayed M, Borahmah A, Yazdani A, Sultan A, Mossad A, Rawdhan H. Assessment of clinical characteristics and mortality-associated factors in COVID-19 Critical cases in Kuwait. medRxiv 2020. DOI: 10.1101/2020.06.17.20134007.

14. De Spiegeleer A, Bronselaer A, Teo JT, et al. The Effects of ARBs, ACEis, and Statins on Clinical Outcomes of COVID-19 Infection Among Nursing Home Residents. J Am Med Dir Assoc 2020;21(7):909–14.

15. Dreher M, Kersten A, Bickenbach J, et al. The Characteristics of 50 Hospitalized COVID-19 Patients With and Without ARDS. Deutsches Arzteblatt International 2020;117(16):271–8.

16. Grasselli G, Greco M, Zanella A, et al. Risk Factors Associated With Mortality Among Patients With COVID-19 in Intensive Care Units in Lombardy, Italy. JAMA Intern Med 2020;180(10):1345–1355. DOI: 10.1001/jamainternmed.2020.3539.

17. Tan WYT, Young BE, Lye DC, Chew DEK, Dalan R. Statin use is associated with lower disease severity in COVID-19 infection. Sci Rep 2020;10(1):17458. DOI: 10.1038/s41598-020-74492-0.

18. Yan H, Valdes AM, Vijay A, et al. Role of Drugs Used for Chronic Disease Management on Susceptibility and Severity of COVID-19: A Large Case-Control Study. Clin Pharmacol Ther 2020;108(6):1185–1194. DOI: 10.1002/cpt.2047.

19. Chow R, Im J, Chiu N, et al. The protective association between statins use and adverse outcomes among COVID-19 patients: a systematic review and meta-analysis. PLOS One 2021;16:e0253576. DOI: https://doi.org/10.1101/2021.02.08.21251070.

20. Kim AY, Gandhi RT. COVID-19: Management in hospitalized adults. UpToDate. (https://www.uptodate.com/contents/covid-19-management-in-hospitalized-adults/print?search=statincovid&source=search_result&selectedTitle=1~150&usage_type=default&display_rank=11/37).

21. Kwon S. Thirty years of national health insurance in South Korea: lessons for achieving universal health care coverage. Health Policy and Planning 2009;24(1):63–71. DOI: 10.1093/heapol/czn037.

22. World Health Organization. International Statistical Classification of Diseases and Related Health Problems 10th Revision. (https://icd.who.int/browse10/2019/en).

23. World Health Organization. Laboratory testing for coronavirus disease 2019 (COVID-19) in suspected human cases: interim guidance, 2 March 2020. Geneva: World Health Organization, 2020 2020. (WHO/COVID-19/laboratory/2020.4) (https://apps.who.int/iris/handle/10665/331329).

24. Ministry of Health and Welfare, Republic of Korea. About COVID-19: patient treatment & management.

25. Lee HY, Ahn J, Park J, et al. Beneficial Effect of Statins in COVID-19-Related Outcomes-Brief Report: A National Population-Based Cohort Study. Arterioscler Thromb Vasc Biol 2021;41(3):e175–e182. DOI: 10.1161/atvbaha.120.315551.

26. Zheng Z, Peng F, Xu B, et al. Risk factors of critical & mortal COVID-19 cases: A systematic literature review and meta-analysis. J Infect 2020;81(2):e16–e25. DOI: 10.1016/j.jinf.2020.04.021.

27. Chow R, Prsic EH, Shin HJ. Cost-Effectiveness Analysis of Statins for Treatment of Hospitalized COVID-19 Patients. medRxiv 2021:2021.04.29.21256335. DOI: 10.1101/2021.04.29.21256335.

28. Shrank WH, Patrick AR, Brookhart MA. Healthy user and related biases in observational studies of preventive interventions: a primer for physicians. J Gen Intern Med 2011;26(5):546–50. DOI: 10.1007/s11606-010-1609-1.

29. Park EC, Jang SI, Jeon SY, Lee SA, Lee JE, Choi DW. Assessment of level of agreement in disease codes between Health Insurance Claims Data and Medical Records. Health Insurance Review & Assessment Service 2018 (http://www.alio.go.kr/informationResearchView.do?seq=2343982).

